# Preoperative activation of the Renin–Angiotensin system and myocardial injury in noncardiac surgery: Post Hoc Analysis of the SPACE randomised controlled Trial

**DOI:** 10.1101/2024.03.22.24304763

**Authors:** Ana Gutierrez del Arroyo, Tom. E. F. Abbott, Akshaykumar Patel, Salma Begum, Priyanthi Dias, David Brealey, Rupert M. Pearse, Vikas Kapil, Gareth L. Ackland, the SPACE trial investigators

**Author notes:** Correspondence to: Gareth L. Ackland Translational Medicine and Therapeutics William Harvey Research Institute, Queen Mary University of London London EC1M 6BQ United Kingdom.

## Abstract

**Background:** Hypertension therapy in older adults is often suboptimal, in part due to inadequate suppression of the renin-angiotensin-aldosterone system (RAAS). We hypothesised that distinct endotypes of RAAS activation before noncardiac surgery are associated with increased risk of myocardial injury.

**Methods:** This was a pre-specified analysis of a multicentre randomised controlled trial (ISRCTN17251494) which randomised patients ≥60 years undergoing elective non-cardiac surgery to either continue, or stop, RAAS inhibitors (determined by pharmacokinetic profiles). Unsupervised hierarchical cluster analysis identified distinct groups of patients with similar RAAS activation from samples obtained before induction of anesthesia, quantified by enzyme-linked immunoassays for plasma renin, aldosterone, angiotensin-converting enzyme 2 (ACE2) and dipeptidyl peptidase-3 (DPP3). The primary outcome, masked to investigators and participants, was myocardial injury (plasma high-sensitivity troponin-T).

**Results:** We identified three clusters, with similar proportions of RAAS inhibitors randomised to stop/continue. Cluster 1 (n=52; mean age (SD), 75±8 years; 54% female) and cluster 3 (n=25; 75±6 years; 44% female) had higher rates of myocardial injury (23/52 (44%) and 13/25 (52%), respectively), compared with 51/164 (31.1%) in cluster 2 (n=153; 70±6 years; 46% female; odds ratio:1.95, 95% CI:1.12-3.39, p=0.018). Cluster 2 was characterized by lower NT-proBNP (mean difference, 698pg.ml^-1^, 95% CI, 576-820) and higher renin (mean difference:350pg.ml^-1^, 95% CI:123-577), compared with clusters 1 and 3 with the higher rate of myocardial injury.

**Conclusion:** Effective preoperative RAAS inhibition is associated with lower risk of myocardial injury before non-cardiac surgery, independent of stopping/continuing RAAS inhibitors before surgery.

---

Despite advances in perioperative care, postoperative complications still lead to delayed recovery and accelerated mortality even after discharge from hospital.^1,2^ The complexity of perioperative pathophysiology necessitates, in part, a patient-specific approach to understand perioperative pathophysiology in more depth, thereby identifying opportunities to reduce the burden of postoperative complications.^3^

Surgical patients most at risk of sustaining myocardial injury, and subsequent complications,^4^ after non-cardiac surgery^5,6^ are commonly prescribed renin-angiotension system inhibitors (RAASi) for cardiometabolic disease and hypertension.^7,8^ Hypertension therapy in older adults is often suboptimal,^9^ which may impact on cardiac complications after non-cardiac surgery by failing to reduce activation of the renin-angiotensin system and inflammation.^10^ The non-canonical axis of the renin–angiotensin system counteracts the deleterious effects of angiotensin II, largely through angiotensin-converting enzyme-2 (ACE2).^11^ As a homolog of ACE, ACE2 converts angiotensin II into the organ protective mediator ang1-7. Dipeptidyl peptidase 3 (DPP3) also positively regulates the RAAS pathway by degrading circulating angiotensin II.^11^ Accordingly, inadequate treatment with RAASi may predispose noncardiac surgical patients to organ injury by failing to reduce aldosterone through the negative feedback effect of angiotensin II on renin release and/or maintaining cellular protection via the counter-regulatory RAAS.

In this pre-specified sub study of the SPACE multicentre, randomized trial of stopping or continuing RASi according to their individual pharmacokinetic profile before noncardiac surgery,^12^ we hypothesized that endotypes of RAAS activation may predispose patients to myocardial injury within 48h of surgery – independently of RAASi cessation/continuation.

## Methods

### Study design and participants

The trial was conducted in accordance with International Council for Harmonisation (ICH) Good Clinical Practice Guidelines and applicable laws and regulations. The study protocol was approved by London research ethics committee (16/LO/1495) and the Medicines and Healthcare products Regulatory Agency (UK). Each patient provided written informed consent. The SPACE trial was reported in accordance with CONSORT guidelines and registered publicly (ISRCTN17251494). From 07/31/2017 to 10/01/2021, the Stopping Perioperative ACE-inhibitors/ARBs (SPACE) phase 2a trial (EudraCT:2016-004141-90) randomised adults aged ≥60 years to either continue or stop their RASi according to the individual pharmacokinetics of each drug. Adults prescribed RAS inhibitors aged ≥60 years, with American Society of Anesthesiologists physical status grade 3 or above undergoing elective major surgery requiring general anaesthesia lasting longer than 120 min were eligible. Exclusion criteria included current participation in any other interventional clinical trials and myocardial infarction within the 3 months preceding surgery. We did not exclude patients taking ACE-I/ARB for left ventricular dysfunction (ejection fraction <50%).

### Randomisation and masking

Randomization was performed centrally, with minimization by centre, RAS inhibitor and type of surgery. After randomization, participants received confirmation of which treatment group they have been allocated to and reminded of their randomized allocation by daily telephone call and/or text message, or in person if they were in hospital. All laboratory personnel undertaking troponin measurements and ELISAs were masked to study details and trial allocation.

### Perioperative drug management

Renin–angiotensin system inhibitors were restarted after surgery on the morning of post-operative day 2, in accord with recommendations by the ESC guidelines.^13^ RAS inhibitors were recommenced at the same dose as that prescribed before surgery. Resumption of RAS inhibitor therapy was delayed on post-operative day 2 if systolic blood pressure was <90 mmHg in the preceding 12 h, vasoactive therapy was required to maintain blood pressure and/or if acute kidney injury had been sustained.^14^ Usual peri-operative practice was delivered in accord with recent ESC peri-operative guidelines.^13^

### Primary and secondary endpoints

The primary endpoint was myocardial injury, defined by Troponin-T levels (Roche) using the VISION study thresholds.^1^ As recommended by the Standardized Endpoints Consensus guidelines for cardiovascular outcomes in perioperative care,^15^ major adverse cardiovascular events and length of hospital stay are also reported.

### Explanatory variables

Blood samples collected before the induction of anaesthesia and on the morning 24 and 48 h after surgery. Batched analyses of plasma samples stored at -80°C were undertaken for troponin (Doctor’s Laboratory, London, UK) and RAAS components. From samples obtained on the day of surgery before induction of anaesthesia, we quantified renin (BMS2271, Invitrogen, Carlsbad, USA), aldosterone (ADI-900-173, Enzo, Farmingdale, USA) angiotensin-converting enzyme 2 (ACE2; DY33-05, R+D Systems, Abingdon, UK), dipeptidyl peptidase-3 (DPP3; E3112Hu, Bioassay Technology Lab, Shanghai, China) and NT-proBNP (ab263877, Abcam, Cambridge UK) using established enzyme-linked immunoassays performed by personnel masked to study details (Tecan plate reader). Standard curves for each ELISA exceeded R^2^ values >0.99 (Supplementary data).

### Statistical analysis

A statistical analysis plan was published before biochemical analyses were undertaken (https://www.qmul.ac.uk/ccpmg/sops--saps/statistical-analysis-plans-saps). We used cluster analysis to identify RAAS endotypes, a data-reduction technique designed to uncover subgroups of observations within a dataset. A cluster is defined as a group of observations that are more similar to each other than they are to the observations in other groups. This has been used in cardiovascular medicine to identify heterogeneity in pathophysiological states ranging from heart failure,^16^ left ventricular dysfunction,^17^ to echocardiographic phenotyping.^18^ Optimal cluster number was determined using NbClust (R4.3.2; Supplementary data).^19^ We used unsupervised hierarchical cluster analysis to generate homogeneous groups of patients by reducing the preoperative plasma values for renin, aldosterone, ACE2, DPP3 and NT-proBNP. Primary and secondary outcomes were analyzed by Fishers exact test and log-rank test, as appropriate. Maximal troponin and RAAS mediator concentrations were analyzed by one-way analysis of variance, with post-hoc Tukey-Kramer testing. Analyses were performed using NCSS 2023 (Kaysville, Utah, USA).

## Results

### Baseline characteristics

Cluster analysis distinguished three groups of patients with similar biochemical characteristics of RAAS activation before surgery (Figure 1). Patients within each cluster underwent similar types of surgery and were prescribed similar proportions of ACE inhibitors/ARBs before surgery (Table 1). The proportion of RASi stopped/continued before surgery by random allocation, were also similar. Cluster 3, who received more anti-hypertensive medications, also developed markedly higher systolic and diastolic blood pressure after randomisation compared to clusters 1 and 2 (Figure 2).

**Figure 1.**
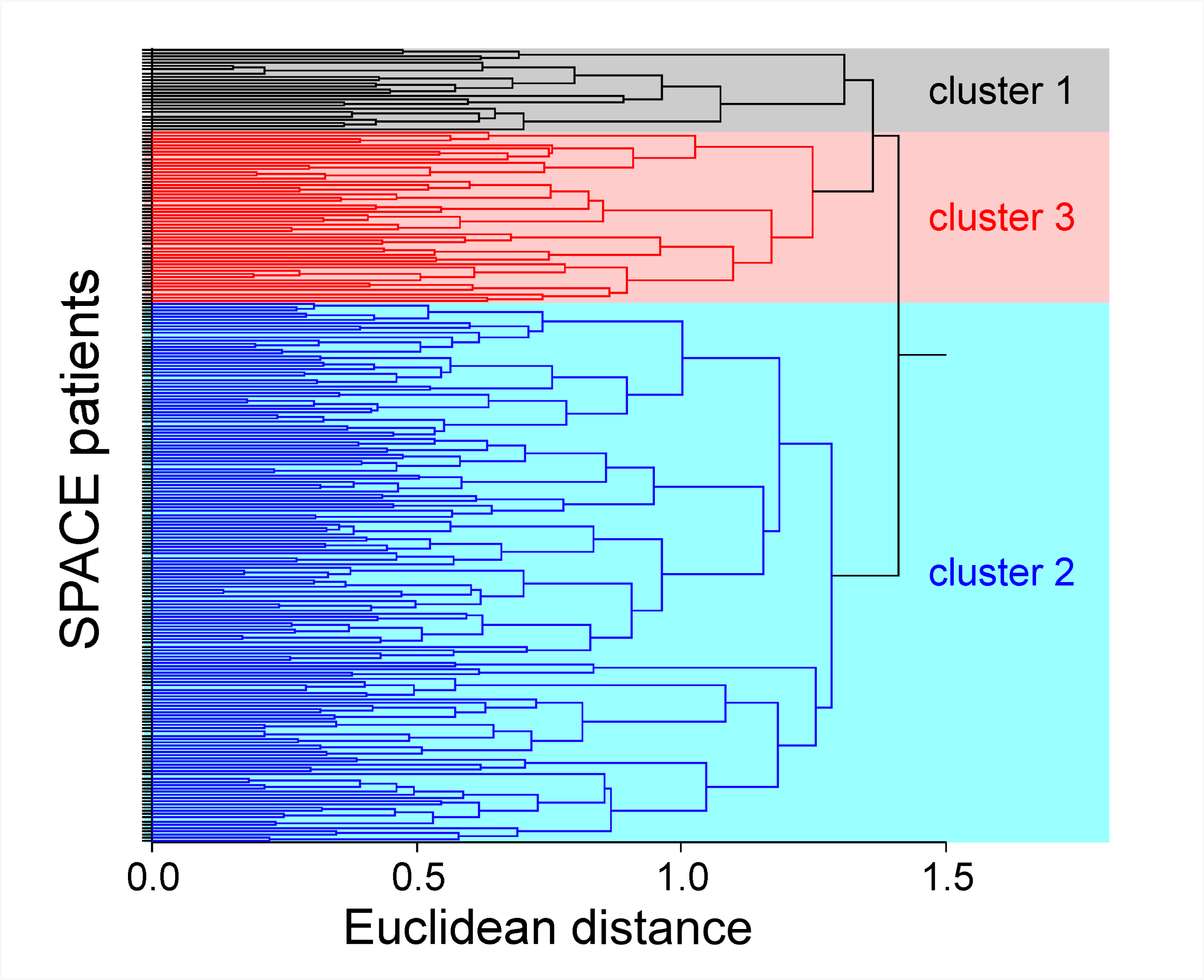
Unsupervised hierarchial cluster analysis. A. Clustering of preoperative RAAS characteristics, determined by unsupervised hierarchial cluster analysis after determination of optimal cluster number by NBClust.

**Table 1.** Participant characteristics by cluster. Data are n (%), mean (SD) or median (Interquartile range); COPD, chronic obstructive pulmonary disease.

|  | **Cluster 1** | **Cluster 2** | **Cluster 3** |
| --- | --- | --- | --- |
| n | 52 | 164 | 25 |
| Female (n; %) | 28 (54%) | 75 (46%) | 11 (44%) |
| Age | 75 (8) | 70 (6) | 75 (6) |
| Haemoglobin before surgery | 127 (24) | 134 (14) | 128 (30) |
| Creatinine | 91 (32) | 87 (29) | 101 (56) |
| ACEi | 33 (63%) | 101 (62%) | 13 (52%) |
| ARBs | 19 (37%) | 63 (38%) | 12 (48%) |
| Randomised to stop ACEi/ARB (n;%) | 24 (46%) | 84 (51%) | 12 (48%) |
| Extra-abdominal surgery | 42 (81%) | 134 (82%) | 18 (72%) |
| COPD | 3 (6%) | 26 (16%) | 0 |
| Asthma | 6 (12%) | 20 (12%) | 2 (8%) |
| Diabetes mellitus | 17 (33%) | 50 (30%) | 6 (24%) |
| Heart failure | 4 (8%) | 11 (7%) | 1 (4%) |
| Cancer | 15 (29%) | 38 (23%) | 8 (32%) |
| Stroke | 2 (4%) | 13 (8%) | 3 (12%) |
| Peripheral vascular disease | 2 (4%) | 11 (7%) | 2 (8%) |
| Hypertension | 51 (98%) | 160 (98%) | 23 (92%) |
| Smoker | 2 (4%) | 16 (10%) | 2 (8%) |
| Number of antihypertensives (1:2:3:4) | 14:20:13:5 | 50:75:31:8 | 5:9:10:1 |
| ≥ 3 antihypertensives inc.RASi (n; %) | 13 (25%) | 39 (24%) | 11 (44%) |
| Beta blocker | 21 (40%) | 42 (26%) | 10 (40%) |
| Calcium channel blocker | 18 (35%) | 60 (37%) | 7 (28%) |
| Doxazosin | 6 (12%) | 13 (8%) | 4 (16%) |
| Diuretic | 16 (31%) | 46 (28%) | 11 (44%) |
| Statin | 33 (63%) | 122 (74%) | 16 (64%) |
| Nitrate | 8 (15%) | 14 (9%) | 3 (12%) |
| Anti-platelet | 18 (35%) | 62 (38%) | 8 (32%) |

**Figure 2.**
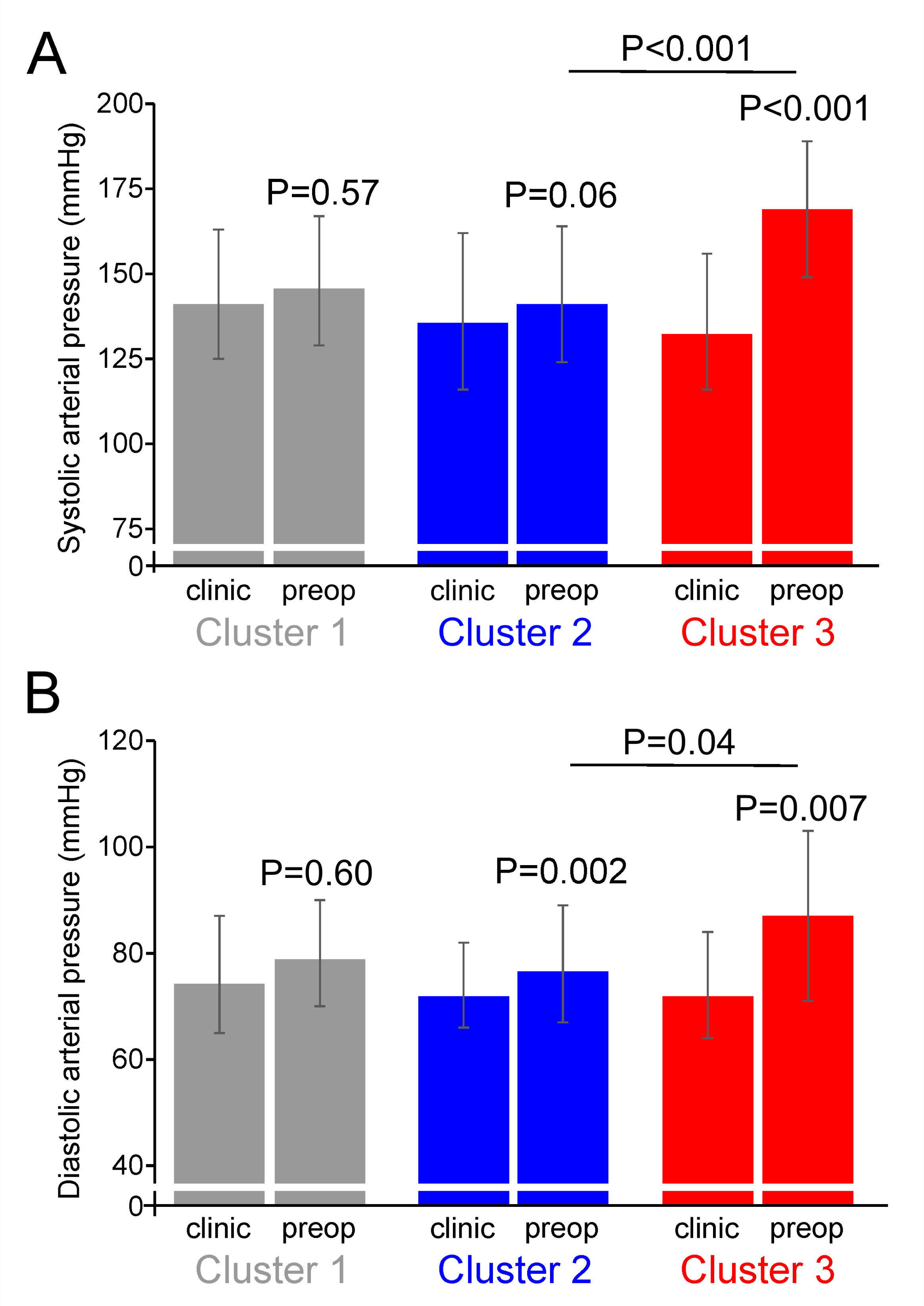
Blood pressure changes before and after randomisation in each cluster. A. Systolic blood pressure, in preoperative clinic and on the day of surgery for each cluster identified by unsupervised cluster analysis. B. Diastolic blood pressure, in preoperative clinic and on the day of surgery for each cluster identified by unsupervised cluster analysis. P values refer to within group post-hoc Tukey Kramer comparisons, plus comparison between clusters 2 and 3.

### Primary outcome: myocardial injury after surgery

For the primary outcome, clusters 1 and 3 had higher rates of myocardial injury 36 of 77 (47%) patients with elevated troponin after surgery (Figure 3A), compared to 51 of 164 patients (31.1%) in cluster 2 (odds ratio, 1.95; 95% CI, 1.12-3.39; p=0.018). A sensitivity analysis excluding beta-blockers (which alter the aldosterone/renin ratio)^20^ showed similar results (odds ratio, 2.09; 95% CI, 1.04-4.21; p=0.038, Supplementary data).

**Figure 3.**
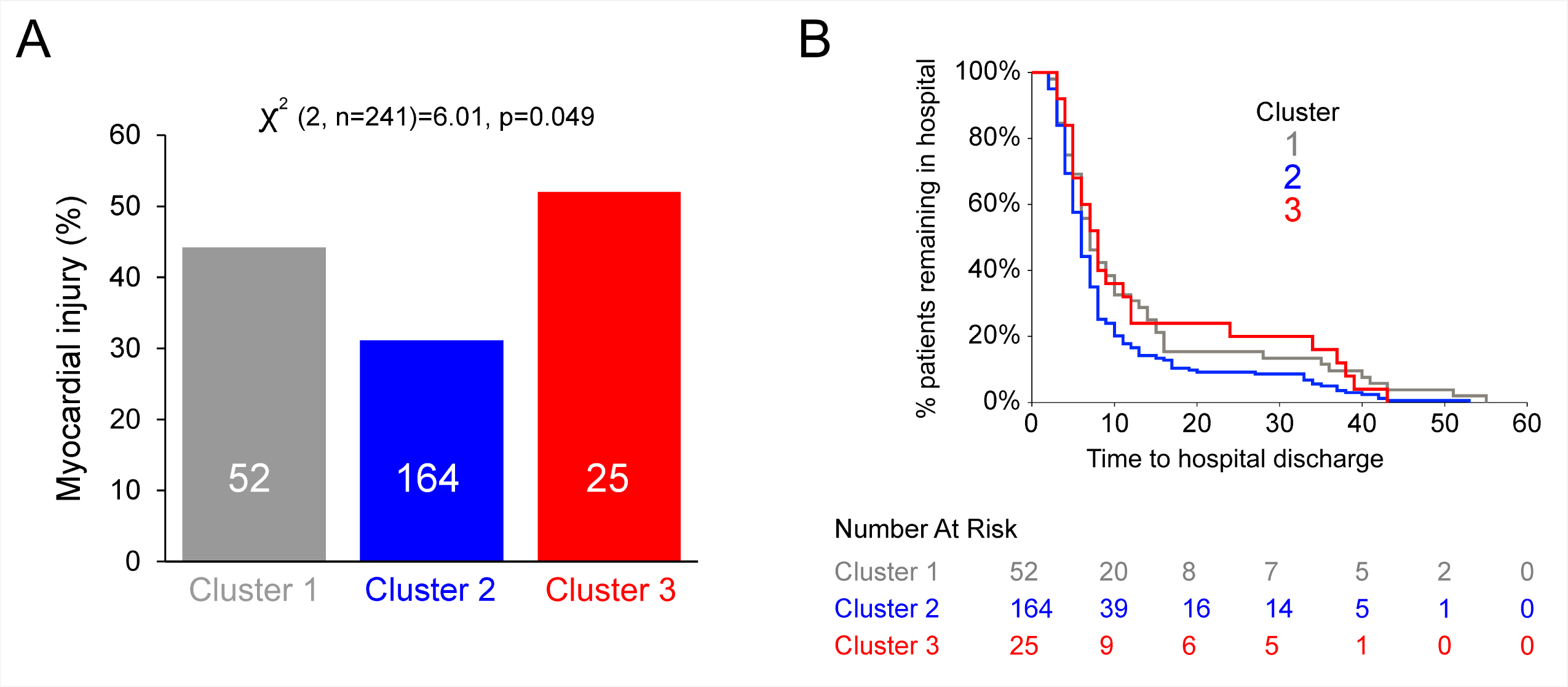
Primary outcome-myocardial injury. A. Incidence of myocardial injury in each cluster. Numbers of patients per cluster are shown within each bar. B. Length of hospital stay, illustrated by time to hospital discharge in Kaplan-Meier plot for each cluster. Patients in cluster 2 had a shorter hospital stay (hazard ratio, 0.73; 95% CI, 0.56-0.95); p=0.012 by log-rank test).

### Secondary outcomes

Mean peak troponin levels were 5ng.l^-1^ lower (95% CI, 1-9) in cluster 2 patients, compared to clusters 1 and 3 (p<0.001). For the entire cohort, myocardial injury was associated with a 3 day (95%CI, 1-5) prolonged hospital stay. Patients in cluster 2 had a shorter hospital stay (hazard ratio, 0.73; 95% CI, 0.56-0.95); p=0.012, by log-rank test; Figure 3B). Clinically defined cardiovascular complications were similar across each cluster (Table 1).

### RAAS characteristics of each cluster

Cluster 2, with the lowest rate of myocardial injury, was characterized by lower NT-proBNP (mean difference, 698pg.ml^-1^ (95% CI, 576-820; Figure 4A) and higher renin (mean difference, 350pg.ml^-1^ (95% CI, 123-577); Figure 4B), compared with clusters 1 and 3 that had higher rates of myocardial injury. Cluster 3 had higher plasma aldosterone (mean difference, 196pg.ml^-1^, 95% CI, 30-362; Figure 4C, D) and ACE2 (mean difference, 3.8ƞg.ml^-1^, 95% CI,1.0-6.6), compared to clusters 1 and 2 (Figure 4E). DPP3 values were similar across each cluster (Figure 4F).

**Figure 4.**
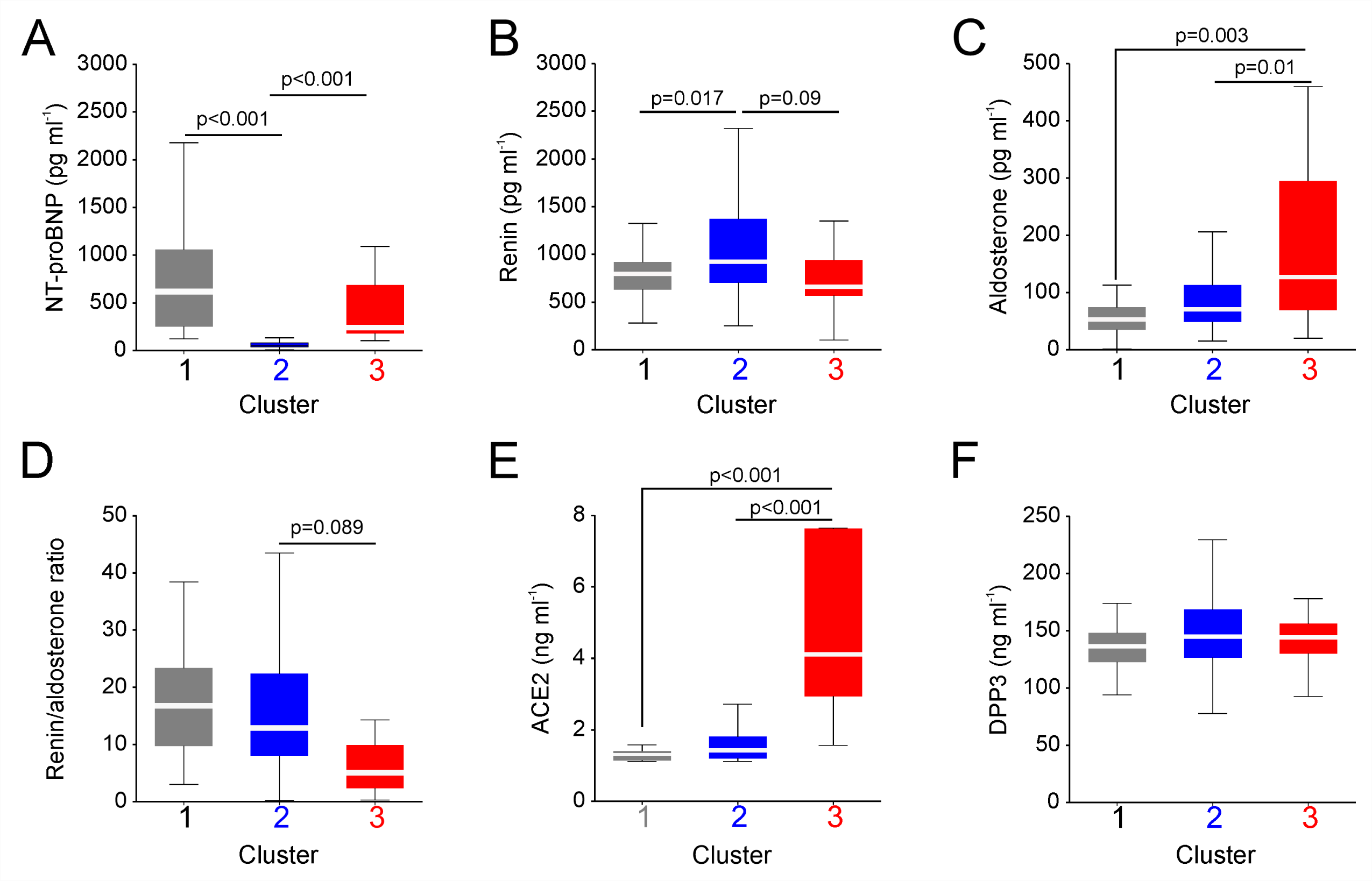
RAS endotypes-plasma levels of RAAS mediators. A. NT-proBNP. B. Renin. C. Aldosterone. D. Renin/aldosterone ratio. E. ACE2. F. DPP-3 All data are median (IQR). P values refer to between group comparisons, as derived from post-hoc Tukey Kramer comparisons.

## Discussion

This pre-defined sub-study of the SPACE RCT found that patients with a preoperative RAAS endotype indicative of adequate RAAS inhibition and/or cardiovascular therapy had a substantially lower risk of sustaining myocardial injury after non-cardiac surgery. These findings were independent of whether the patients enrolled into SPACE were randomised to continue or stop their ACE inhibitors or ARBs. Patients at lowest risk of sustaining myocardial injury had substantially lower preoperative NT-proBNP. Taken together, these data indicate that chronic, under-treated RAAS activation contributes to perioperative myocardial injury (summarised in Figure 5), and is consistent with the findings of the main SPACE trial where stopping ACEi/ARBs was associated with higher rates of myocardial injury.

**Figure 5.**
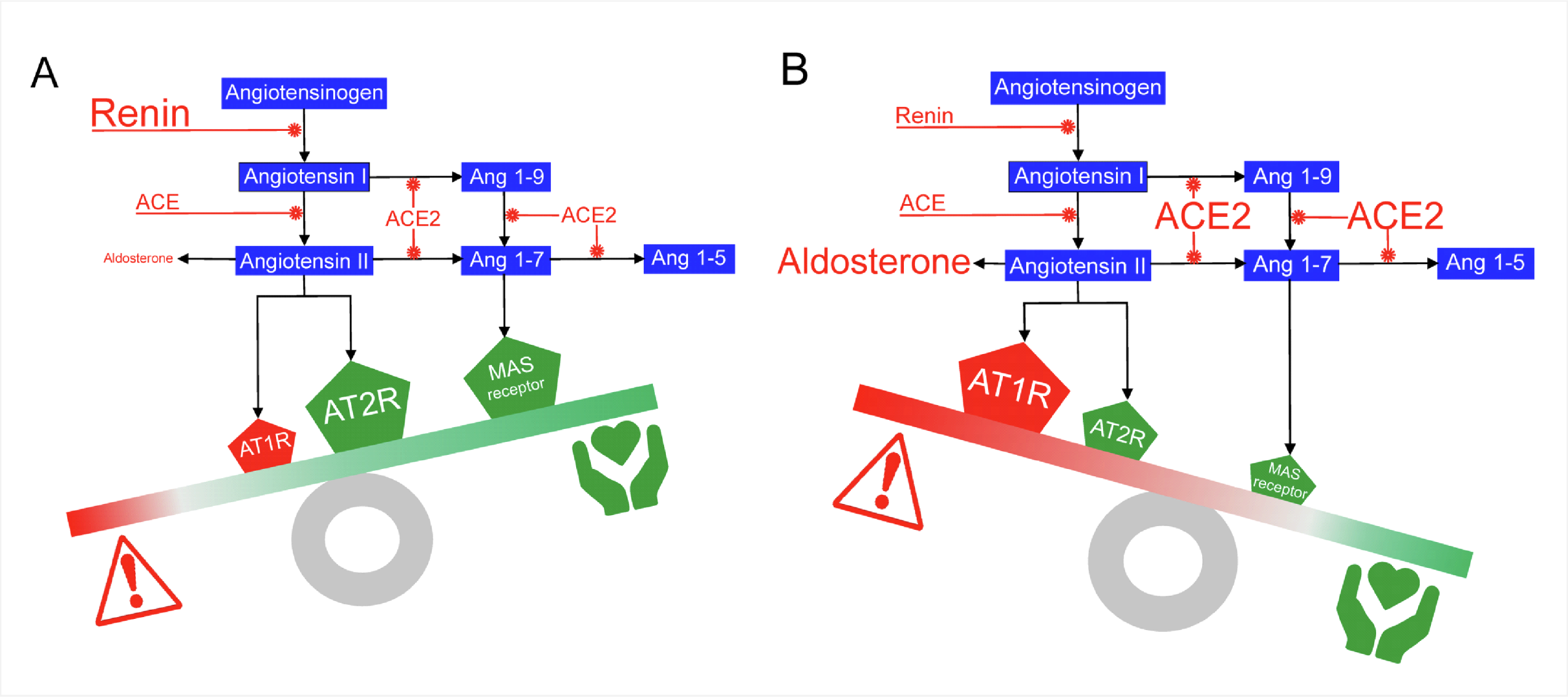
Preoperative RAS endotypes and factors altering risk of myocardial injury. A. Effective ACE inhibition and/or angiotensin-II blockade promote cardioprotective signalling pathways via reduction in ROS/inflammation. B. Lack of effective RAAS suppression leads to excess aldosterone and ACE2 expression, promoting diastolic and endothelial dysfunction and worse perioperative outcomes.

The two clusters with higher rates of myocardial injury were characterised by distinct clinical features and biochemical indicators of RAAS activation that are mechanistically linked to cardiomyocyte damage. Cluster 3, comprising ∼10% patients, had marked elevation in plasma aldosterone, a key mediator in the pathogenesis of heart failure and consistent with primary aldosteronism evident in 8–13% non-selected patients with essential hypertension.^21^ Aldosterone increases cardiac inflammation, fibrosis, and myocardial hypertrophy through numerous mechanisms.^22^ Aldosterone impairs endothelial function by reducing nitric oxide production in endothelial cells through the inhibition of endothelial nitric oxide synthase activity.^23^ Aldosterone reduces glucose-6-phosphate dehydrogenase in endothelial cells, promoting oxidative stress and reduced nitric oxide bioavailability.^24^ Aldosterone also increases the expression of adhesion molecules in both endothelial and coronary artery smooth muscle cells,^25^ fuelling inflammation through injurious leukocyte-endothelium crosstalk and leading to consequent extravasation.^26^

Patients with primary aldosteronism have structural and functional changes characteristic of diastolic dysfunction, with thickened ventricular walls, decreased mitral early peak velocity and 20% lower peak myocardial early velocity.^27^ These mirror patients with preserved ejection fraction heart failure, who experience high rates of postoperative morbidity and mortality after non-cardiac surgery. From one database study involving 153,771 patients with preserved ejection fraction heart failure, 40% sustained cardiopulmonary morbidity, while non-cardiovascular complications including acute kindey injury and sepsis were recorded in 55% patients.^28^ These complications were associated with a staggeringly high 5% in-hospital mortality rate and 20% 30-day hospital readmission rate.

As a clinical consequence, high aldosterone levels are very likely associated with enhanced risk of cardiovascular events and mortality, especially when aldosterone secretion is inappropriate for renin levels and sodium intake. Cluster 3 also had markedly raised plasma ACE2 concentration, which is associated with increased risk of major cardiovascular events.^29^ Higher plasma ACE2 concentrations have been reported in patients with primary aldosteronism.^30^ Patients in cluster 3 also demonstrated marked increases in blood pressure on the day of surgery, compared to their preoperative values. Comprising ∼10% of our study population, these data are consistent with a prevalence of resistant hypertension of 10%–11% in the UK in among apparently compliant patients aged 70 and older.^31^

In cluster 1, the combination of lower renin and elevated NT-proBNP was associated with similarly higher rates of myocardial injury to cluster 3. The significantly lower plasma renin levels in contrast to the higher levels displayed in the relatively cardioprotected cluster 2 are compatible with ineffective RAAS inhibition. ACEIs and ARBs inhibit the formation and action of ANG-II, respectively, and consequently suppress the negative feedback inhibition of renin release by ANG-II.^32^ Accordingly, a compensatory increase in plasma renin concentration occurs accompanied by an increase in plasma renin activity.^33^ Inhibition of ACE also results in increased ANG-I levels, which may be converted to ANG-II via non– ACE-dependent pathways through reactive hyper-reninaemia.^34^ Blockade of angiotensin II stimulates renin synthesis and release indirectly through the action of ligands that activate the cAMP/PKA pathway in a Gsα-dependent fashion, including catecholamines, prostaglandins, and nitric oxide. The ACE inhbitiors captopril and quinaprilate and the ARB candesartan increase plasma renin concentration 20-40 fold above basal levels in wild-type mice, but do not alter plasma renin concentrations in Gsα-deficient mice.^35^ Taken together, our trial data suggest that ineffective RAS blockade over time failed to increase the plasma concentration of renin through the negative feedback effect of angiotensin II on renin release. Moreover, our data suggest that the interpretation of perioperative trials exploring the management of RAS inhibitors should be refined by the use of relevant cardio-endocrine biomarkers such as NT-proBNP.

Strengths of our study are that all assays were batch analysed by investigators masked to clinical and trial data. Troponin thresholds used to define myocardial injury are consistent with international guidelines.^5,36^ A limitation was not establishing the etiology of raised troponin by electrocardiographic criteria, although elevated troponin after surgery is associated with poorer outcomes regardless of etiology.^4^ Routine preoperative echocardiography may have added further insights. Given the equal distribution of stopping/continuing RASi in each cluster, sampling before and after randomisation is likely inconsequential. The measurement of additional RAAS components, including Mas and Ang1-7 may have added additional insights. Sampling before stopping RAS inhibitors may also have been more instructive but due to the logistic constraints of patients frequently being located far from referring hospitals, this was impractical. Given the magnitude of differences between the clusters, and the equal proportions of patients randomised to stop or continue RAS inhibitors in each cluster, this does not appear likely to have been a major confounder in interpretation of the results.

In conclusion, preoperative RAAS activation in patients undergoing non-cardiac surgery is associated with myocardial injury. Continuation of RASi and/or preoperative intensification of RAAS inhibition could reducing perioperative myocardial injury.

## Data Availability

Available on request from senior author.

## Sources of Funding

G.L.A. was supported by the National Institute for Academic Anaesthesia (British Oxygen Company research chair grant); NIHR Advanced Fellowship [NIHR300097], and a British Heart Foundation Programme grants (RG/14/4/30736; RG/19/5/34463). T.E.F.A. and R.M.P. were supported by NIHR.

## Disclosures

GLA, RMP DB, TEFA receive support from *NIHR* (National Institute for Health and Care Research). The remaining authors have no disclosures to report.

## Acknowledgments

We thank the SPACE trial patients and their families, trial investigators and support staff, TSC and DMEC independent Committee members.

## Notes

### Competing Interest Statement

The authors have declared no competing interest.

### Clinical Trial

ISRCTN17251494). From 07/31/2017 to 10/01/2021, the Stopping Perioperative ACE-inhibitors/ARBs (SPACE) phase 2a trial (EudraCT:2016-004141-90)

### Clinical Protocols

https://www.qmul.ac.uk/ccpmg/sops--saps/statistical-analysis-plans-saps

### Author Declarations

London research ethics committee (16/LO/1495) and the Medicines and Healthcare products Regulatory Agency (UK).

